# A study of clinical outcomes and prognostic factors associated with invasive mechanical ventilation of patients in non-ICU settings: A systematic review and meta-analysis

**DOI:** 10.1101/2021.04.04.21254885

**Authors:** Shubham Agarwal, Animesh Ray, Abhishek Anand, Neha Chopra, Ananthu Narayan, Vishakh Keri, Neha Rastogi, Debarchan Roy, Ranveer Singh Jadon, Naval K Vikram

## Abstract

There is paucity of evidence related to mechanical ventilation in the general ward setting. We aimed to study the clinico-etiological profile, outcomes and prognostic factors of patients receiving invasive mechanical ventilation in non-ICU (ward) setting, and compare these parameters with that of patients in the ICU, wherever it was reported. A systematic review and meta-analysis was done on articles published till June 2020. Two authors independently extracted the data. The study population included patients who received mechanical ventilation in ward setting. Fourteen studies reporting on 20833 patients were included (20252 exclusively ventilated in ward), with most of the studies being from Israel, USA, Japan and Taiwan. Risk of bias was estimated using the Newcastle-Ottawa Scale for observational studies, and was found to be low. Most common reason for intubation was respiratory illness. Most common variables predicting mortality were prognostic scores like APACHE-II and Acute Physiology Score (APS). Pooled mortality rate in ward across 6 studies was 0.72 (95% CI 0.69-0.74) with no heterogeneity among these 6 studies (I2=0.0). Mortality rate varied significantly with study population characteristics, and was lower among patients being weaned in ward. A major limitation of our study was the paucity of studies and significant heterogeneity among existing studies, with respect to outcomes like duration of ventilation, hospital stay, rates of complications, and prognostic factors. This systematic review and meta-analysis found that mortality among patients receiving invasive mechanical ventilation in ward settings remains high. Data regarding other outcomes and prognostic factors predicting mortality was very heterogeneous highlighting the need for future studies concentrating specifically on these aspects.

Systematic review registration: PROSPERO 2020 (CRD42020166775)

## Introduction

Invasive mechanical ventilation has been classically thought to be exclusive to the ICU (Intensive Care Unit) setting. Key features that make an ICU different from a general ward are the frequency and methods of monitoring, staff-patient ratio, training of staff and presence of respiratory care intensivists and physiotherapists. However, with the rising burden of patients on a struggling health-care system, especially in wake of the COVID-19 pandemic^1^, there is an unmet need for invasive mechanical ventilation in the non-ICU setting. Various levels of step-down units have been available for management of such critical patients once they are discharged from the ICU, including high dependency units (HDUs), weaning units (WUs) and respiratory care wards (RCWs). Many countries, especially the lower-middle income countries (LMIC), do not have the resources for such an elaborate hierarchical setup. There is a huge shortage of ICU beds in these settings and deserving patients are often denied ICU beds due to this reason. The necessity for invasive mechanical ventilation in the general medicine ward is now no longer limited to LMIC nations but is being increasing recognised in all nations^2^. In such general medicine wards, often, both critical intubated and stable non-intubated patients are managed by the same team of doctors and paramedical staff, with the same staff-patient ratio. There is often no availability of respiratory care intensivists and physiotherapists to guide the staff in the ward. Such a scenario is common in many countries and is an essential part of their health care system. There is paucity of data on clinical outcomes and prognostic factors associated with mechanical ventilation in the ward setting. We understand that invasive mechanical ventilation in the general ward is necessary, but we do not know how effective and beneficial it is, especially when compared to ICU. There has not been a systematic review (SR) till date that has summarised and analysed all the available literature on invasive mechanical ventilation in the general ward. Health care sector today has been burdened by the COVID-19 pandemic caused by the SARS CoV-2 virus, witnessing a sudden spurt in the number of patients being mechanically ventilated. ICU beds are full round-the-clock, and mechanical ventilation in general wards is now a necessity in most countries. Through this systematic review, we intend to shed some light on the clinico-etiological profile, outcomes and prognostic factors associated with invasive mechanical ventilation in the ward setting, and compare these parameters with the ICU setting.

## Patients and methods

We conducted a systematic review and meta-analysis on the clinical outcomes and prognostic factors associated with invasive mechanical ventilation in the general wards. It was registered in PROSPERO (CRD42020166775) and reported in accordance with Preferred Reporting Items for Systematic Reviews and Meta-analyses (PRISMA) check list.

### Search strategy

We first searched the PubMed and Embase databases for any existing SR on the topic without any language restrictions. We then searched these databases, without date of publication restrictions, till 30^th^ June 2020, with the indexed search terms:

### PubMed database

(invasive) mechanical ventilation AND (general) ward NOT ICU, mechanical ventilation AND general ward vs ICU, mechanical ventilation AND medicine ward NOT noninvasive, mechanical ventilation AND ward vs intensive care unit, mechanical ventilation AND non-ICU.

### Embase database

mechanical ventilation AND ward vs intensive care unit: ab, artificial ventilation AND general ward: ab, artificial ventilation AND non-ICU

The reference lists of the included studies were manually searched to identify other eligible articles which were then considered for inclusion in this review.

### Study selection

The citations after the initial searches were first exported to Zotero. After removing the duplicate citations, SA and AA independently went through the individual abstracts, excluding the articles not relevant to the topic under review. Any disagreement was discussed with another author (AR) and resolved. Full text of the remaining articles was retrieved and considered for inclusion as per the criteria as mentioned below. Full text articles were obtained using the following inclusion criteria:

1. All patients have received invasive mechanical ventilation at some point during the hospital course
2. Patients have stayed in the general ward at some point while being on mechanical ventilation
3. Studies where the entire cohort was not consisting of intubated mechanically ventilated patients were included if the study had a subgroup analysis for the intubated patients whose data could be extracted

Articles in languages other than English were excluded.

### Risk of bias

Assessed using the Newcastle Ottawa Scale for Cohort/observational studies

### Data extraction

Reviewers (SA and AA) independently extracted the data on an MS Excel spreadsheet. All discrepancies were resolved by another reviewer (AR). Data was extracted under the following headings: title, author, journal, year, country of origin, study design, exclusion criteria, number of patients/events, mean age, % males, most common and other diagnoses, mortality rate, scores like APACHE (Acute Physiology and Chronic Health Evaluation), Charlson score, MODS (Multiple Organ Dysfunction Score), APS (Acute Physiology Score) and ISS (Injury Severity Score), blood pressure, vasopressor use, urine output, investigations like haemoglobin, total leucocyte count, serum creatinine, albumin, mean duration of ventilation, mean length of hospital stay, tracheostomy rate, mean ABGs per day and complications like VAP (ventilator associated pneumonia) rate, pressure sore, GI bleed, barotrauma, self extubation rate, re-intubation rates, and factors associated with death or survival.

### Data analysis

Qualitative and quantitative analyses were done on the data obtained from the included studies. Meta-analysis was done to calculate pooled mortality among patients, using a Forest plot analysis with fixed effect (Stata version 12). Other outcomes like duration of hospital stay and duration of mechanical ventilation were reported through qualitative description. Prognostic factors associated with survival or mortality (as per statistical methods used in individual studies) were also described qualitatively.

## Results

Our search strategy found 43 relevant citations which were exported to Zotero. After removing 12 duplicate articles, 31 abstracts were screened. 11 articles were not found to be relevant, and one was in Spanish. The remaining 19 articles were read in full and five were further excluded based on our exclusion criteria (all patients not mechanically ventilated or not ventilated in general ward). Fourteen studies^3,4,5,6,7,8,9,10,11,12,13,14,15,16^ were reviewed, and the data extracted was analysed. A total of 20833 patients were mechanically ventilated in ward across 12 studies ^4,5,7,8,9,10,11,12,13,14,15,16^ of which 20252 were exclusively ventilated in ward. The 2 remaining studies ^3,6^ assessed for rates of events/incidents associated with care of intubated patients^3^, with 182 events reported in ward compared to 79 in ICU, and outcomes of unplanned extubation^6^. On the other hand, 20940 patients were mechanically ventilated in ICU across 4 studies ^9,10,14,16^ with 34 patients exclusively in ICU.

Most of the studies ^9,12,13,14,15^ were from Israel where the issue of non-availability of critical care beds in ICU is a common occurrence, while others were from Japan^3,10^, Taiwan^4,6^, USA^5,7,16^, Hong Kong^8^, Thailand^11^ (Supplementary Appendix Fig. S1). 11 studies were conducted in a single centre, while 3 ^3,10,12^ were multi-centre studies. 10 studies were retrospective record-based studies^3,4,5,6,7,8,10,12,15,16^, and 4 were prospective observational studies^9,11,13,14^. In all 14 studies, the study group received invasive mechanical ventilation at some point of time. Six studies^3,6,9,10,14,16^ had a comparator arm in the ICU while 8 studies ^4,5,7,8,11,12,13,15^ had a single study group. In eight studies^8,9,10,11,12,13,14,15^, patients were ventilated in the ward exclusively during their entire course of mechanical ventilation, while in one study^9^, patients in the ICU group were ventilated in the ICU exclusively.

Exclusion criteria defined individually in the studies included short duration of mechanical ventilation, defined variably as <2 hours^15^, <24h^8^ and <3 days^10^, non-medical indication of admission ^8,9,13,14,15^, formal DNR (do not resuscitate) order^9,11,13,15^ or post CPR (cardio-pulmonary resuscitation)^15^, short life-expectancy^9,11^ or a diagnosis of cancer^10^ or stroke^14^, and prolonged tracheotomy before admission^6,13,15^. The criteria were common to ward and ICU group.

**PRISMA study flowchart**

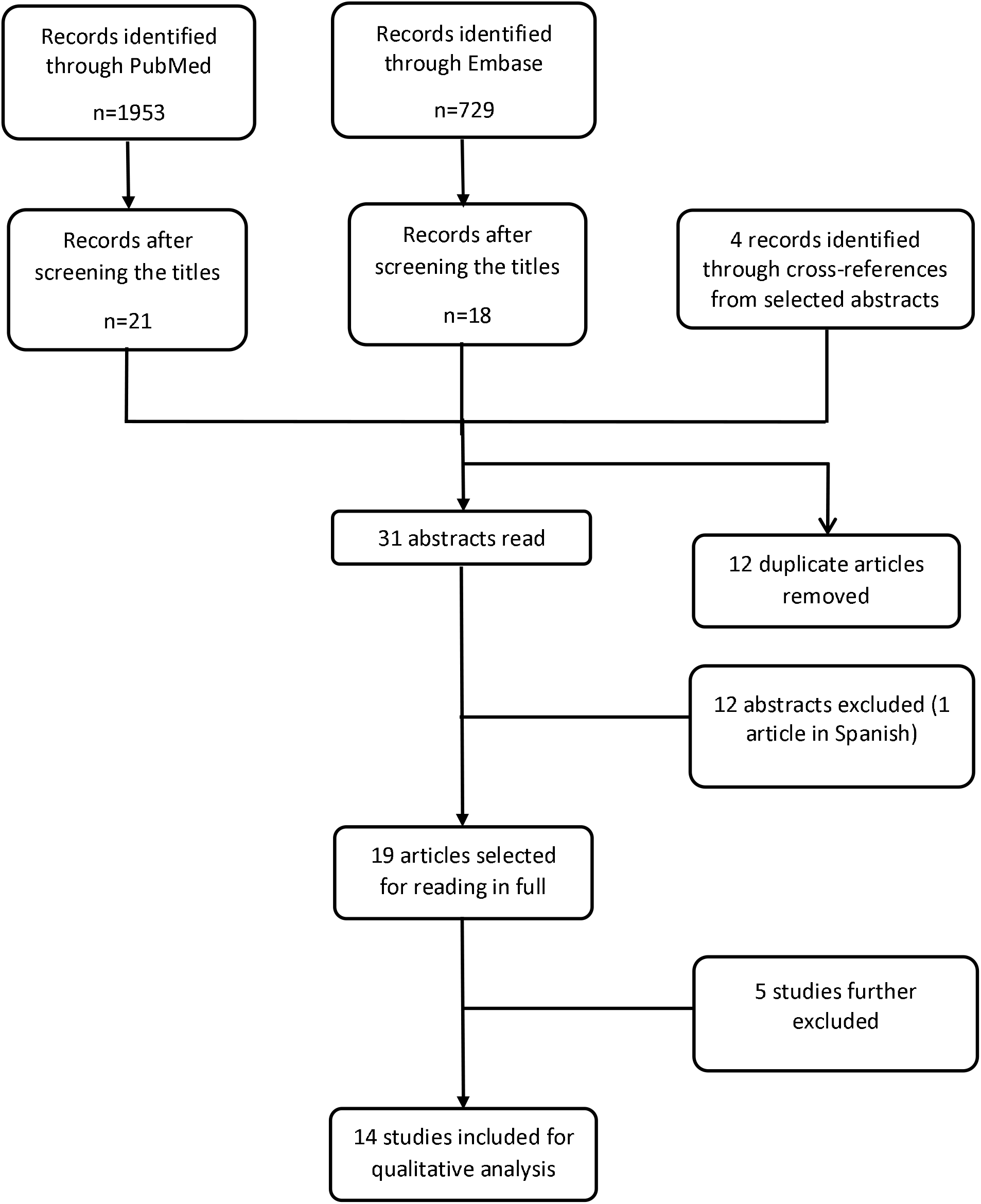

### Quality (Risk of bias) assessment of studies

Quality assessment of 6 studies^4,9,11,13,14,15^ included for analysis of the pooled mortality was assessed (Supplementary Appendix Table S1) using the Newcastle Ottawa Scale for Cohort studies^17^. This was done individually by two authors (SA and AA) and compared. Any discrepancies were resolved after discussion with AR. The scale has parameters to assess both exposed and non-exposed cohorts in a study. However, as none of our observational studies had a non-exposed control arm, few parameters of the tool were not applicable and have been skipped. The final assessment of quality was modified accordingly. All the six studies were rated to be of good quality.

### Clinico-etiological profile

**Table 1:** Baseline characteristics of patients in ward

| Study population | Ward |
| --- | --- |
| Paediatric | 1 study<sup>16</sup> |
| Adult ($\geq 18$ y) | 10 studies<sup>4,5,6,8,9,10,12,13,14,15</sup> |
| Males (%) | Ward |
|  | 11 studies<sup>4,5,6,7,8,9,10,11,13,14,16</sup> |
|  | 11973/19753 (60.6%) |
| Diagnosis | Ward |
| **Cardiovascular** | **3496 (17.2%)** |
| Acute coronary syndrome | 595 |
| Pulmonary oedema/Heart failure | 2561 |
| Arrhythmia | 21 |
| Aortic dissection | 211 |
| Cardiogenic shock | 2 |
| Others/not specified | 106 |
| **Neurological** | **1409 (6.9%)** |
| Ischemic stroke | 65 |
| Intracranial bleed | 1171 |
| Others/not specified | 173 |
| **Respiratory cause** | **4619 (22.7%)** |
| COPD/Asthma | 136 |
| ARDS | 12 |
| Respiratory failure | 1256 |
| Pneumonia | **2970** |
| -Interstitial | 843 |
| -Community acquired | 834 |
| -Hospital acquired | 13 |
| -Aspiration | 1005 |
| -Others/not specified | 275 |
| Others/not specified | 245 |
| Sepsis (non-respiratory) | 682 (3.4%) |
| MODS | 46 (0.2%) |
| Post-operative | 113 (0.6%) |
| Post CPR | 73 (0.4%) |
| Trauma | 76 (0.4%) |
| Uraemia | 29 (0.1%) |
| Metabolic | 10 (0.05%) |
| Neoplastic | 50 (0.2%) |
| Drug overdose | 11 (0.05%) |
| Others/not specified | 9725 (47.8%) |
| **Total** | **20339** |

| Most common diagnosis overall |  |
| --- | --- |
| Neurological | 4 studies<sup>3,4,9,11</sup> |
| Post-operative | 1 study<sup>5</sup> |
| Sepsis (non-pneumonic) | 1 study<sup>14</sup> |
| Respiratory | 3 studies<sup>13,15,16</sup> |
| - Pneumonia | 2 studies<sup>8,11</sup> |
| Heart failure | 1 study<sup>10</sup> |

**Table 2:** Severity and prognostic scores

| Variable | Study | Significant findings* |
| --- | --- | --- |
| APACHE score | Latriano et al<sup>5</sup> | Mean APACHE II score in ward was 16.5±4.8, with higher score in non-survivors (18.2±4.6) as compared to survivors (14.9±4.5). |
|  | Hersch et al<sup>9</sup> | Mean APACHE II score in ward was 27±7. Using multivariate regression, day 1 APACHE II score was found to predict mortality. |
|  | Wongsurakiat et al<sup>11</sup> | APACHE II score of non-survivors (23.3±7.3) was significantly higher than survivors (19.8±5.5). APACHE score >22 was an independent predictor of mortality. |
| Charlson score | Hersch et al<sup>13</sup> | Mean Charlson score was 4 ± 2.2, with no significant difference between survivors and non-survivors. Higher score was not associated with death. |
|  | Lieberman et al<sup>14</sup> | No significant difference between mean score in ward and ICU. Higher Charlson score was an independent predictor of mortality. |
| Acute physiology score (APS) | Hersch et al<sup>13</sup> | Average acute physiology score (APS) among 86 patients in ward was $91.8 \pm 26.7$, with significantly higher score among non-survivors ($98.6 \pm 25.2$) as compared to survivors ($71.8 \pm 20.9$). At APS > 80, probability of death was 5 times higher. |
| | Lieberman et al<sup>14</sup> | Mean acute physiology score (APS) in ward was $15 \pm 5$, with higher APS score being an independent predictor of mortality. |
| Functional status (FIM score) | Lieberman et al<sup>14</sup> | The average total FIM score, motor and cognitive FIM scores in the non-ICU cohort were $75.8 \pm 31.2$, $51.3 \pm 24.1$ and $24.3 \pm 8.8$, respectively. |

\*Findings are statistically significant in individual studies (p values<0.05)

The baseline characteristics of patients in the ICU, distribution of vital parameters, investigations, monitoring and treatment parameters, invasive mechanical ventilation related events have been discussed in the supplementary appendix in tables S3-7, while table S2 gives a distribution of studies based on the main ventilation parameter studied. Supplementary table S8 gives a comparison between significant clinical, monitoring and treatment parameters between ward and ICU.

## Outcomes

### Mortality outcomes

Pooled mortality rate in ward was assessed across six studies^4,9,11,13,14,15^ using a Forest plot analysis with fixed effect. The cumulative incidence of mortality was 0.72 (0.69-0.74) with no heterogeneity among these 6 studies (I^2^=0.0).

**Figure 1:**
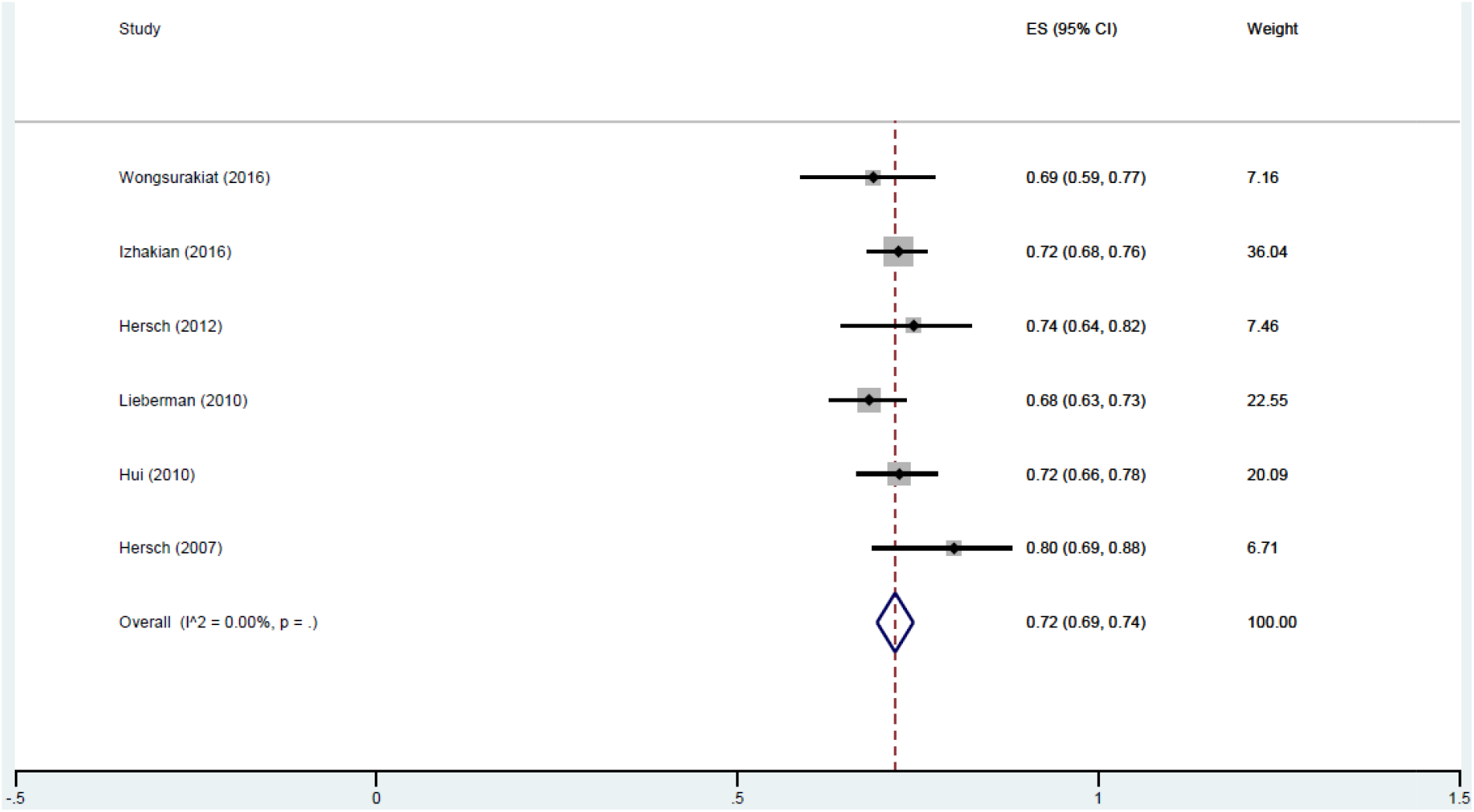
Forest plot analysis showing pooled mortality in ward

Five studies^5,6,7,8,10^ were excluded from the Forest plot as they were significantly contributing to the heterogeneity with very low or high mortality rates. Reasons for their exclusion and heterogeneity have been mentioned in the supplementary appendix (Table S9).

**Table 3:**
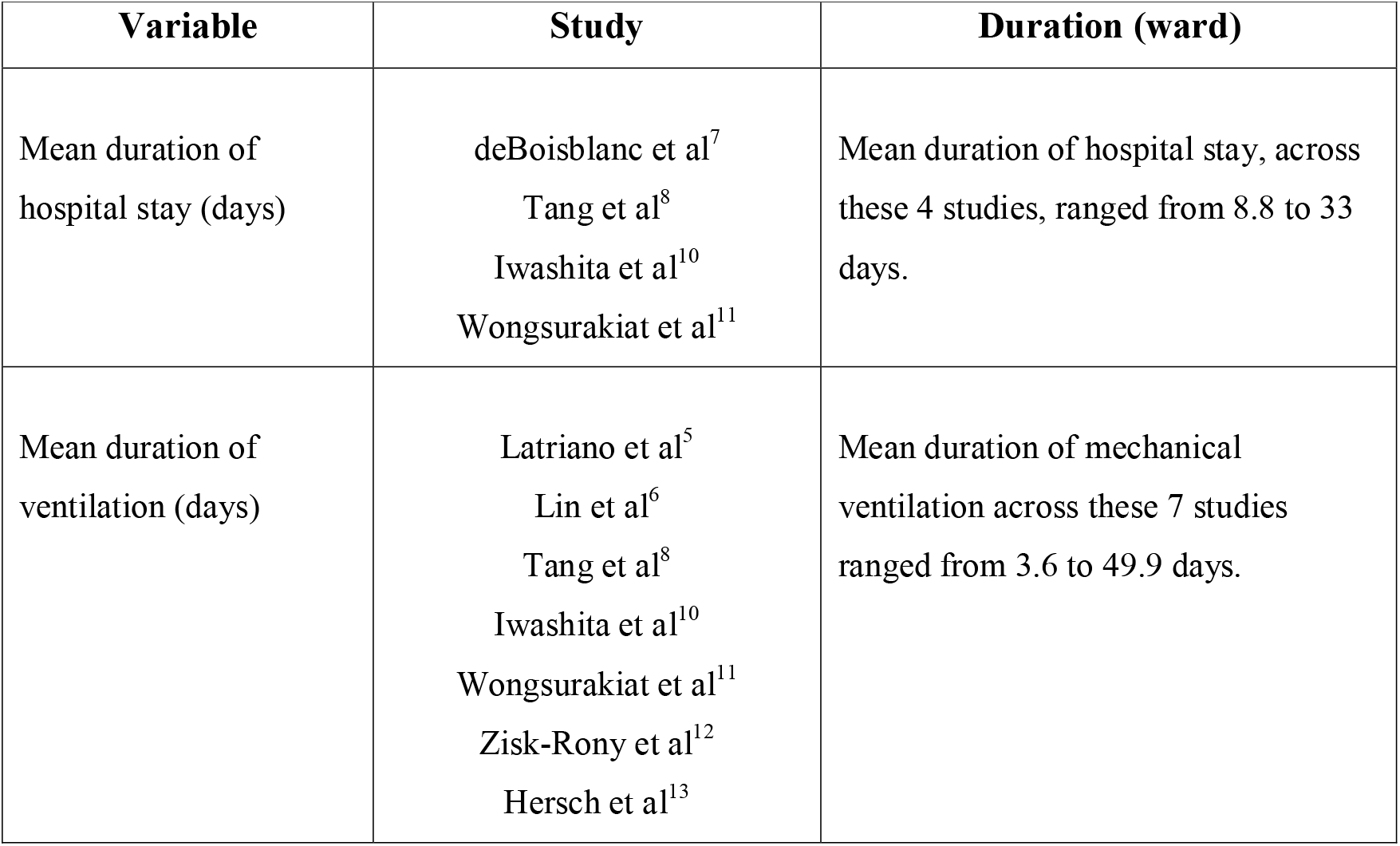
Duration of hospital stay:

**Table 4:** Ventilation related complications:

| Variable | Study | Significant findings<sup>*</sup> |
| --- | --- | --- |
| Need for tracheostomy | Latriano et al<sup>5</sup> | Out of 224 patients, 161 (72%) underwent tracheostomy. |
|  | deBoisblanc et al<sup>7</sup> | All 51 patients were weaned to tracheostomy, of whom 45 (89%) were ultimately weaned to room air. |
|  | Tang et al<sup>8</sup> | Out of 755 patients, 6 (0.8%) underwent tracheostomy. |
| Ventilator associated pneumonia (VAP) | deBoisblanc et al<sup>7</sup> | Out of 51 patients in ward, 22 (43%) developed ventilator associated pneumonia. |
|  | Tang et al<sup>8</sup> | Out of 755 patients in ward, 52 (6.9%) developed ventilator associated pneumonia. |
| Re-intubation | Lin et al<sup>6</sup> | Out of 102 unplanned extubations in ward, 42 patients (41.2%) were re-intubated, while out of 220 unplanned extubations in ICUs, 100 patients (45.5%) were re-intubated. |
|  | Izhakian et al<sup>15</sup> | Out of 437 patients, 43 (9.8%) underwent recurrent intubation, of whom 36 died (84%). |
| Pressure sores, stress ulcer bleed and barotrauma | Tang et al<sup>8</sup> | Out of 755 patients in ward, pressure sores were seen in 57 patients (7.5%), stress ulcer bleeding in 68 (9%) and barotrauma in 4 (0.5%) patients. |

**Table 5:** Factors independently associated with mortality in ward

| Prognostic factor | Study |
| --- | --- |
| Emergency admission<br>Non-cardiac diagnosis<br>Intubation >4d after admission<br>Anaemia at baseline<br>Catecholamine use<br>Renal replacement therapy<br>Bacterial/fungal sepsis | Iwashita et al<sup>10</sup> |
| Higher APACHE-II score | Hersch et al<sup>9</sup><br>Wongsurakiat et al<sup>11</sup> |
| Higher APS component of APACHE-III score | Hersch et al<sup>13</sup><br>Lieberman et al<sup>14</sup> |
| Diagnosis of sepsis<br>Higher Charlson score | Lieberman et al<sup>14</sup> |
| Intubation in ward (after admission)<br>Recurrent intubation | Izhakian et al<sup>15</sup> |

## Discussion

Our systematic review reveals that most of the patients receiving mechanical ventilation in ward settings were males, with mean age ranging from 60-80 years. Only one paediatric study was included. Most of the studies were from Israel, followed by USA and Japan. Most common diagnosis in wards was respiratory illness (including pneumonia),^8,11,13,15,16^ followed by neurological diseases^3,4,9,11^. Mechanical ventilation related outcomes, like mortality rate, duration of mechanical ventilation, total duration of hospital stay, individual complication rates were also assessed. A meta-analysis was done to calculate the pooled mortality rate in ward, across 6 studies^4,9,11,13,14,15^, using a Forest plot analysis with fixed effect. The cumulative incidence of mortality was 0.72 (0.69-0.74) with no heterogeneity among these 6 studies (I^2^=0.0). There were five studies^5,6,7,8,10^ which were excluded due to significant heterogeneity. The duration of hospital stay or of mechanical ventilation, was very variable, depending on the patient demographic and the main parameters being studied. The mean duration of hospital stay ranged from 8.8 to 33 days^7,8,10,11^, while the mean duration of mechanical ventilation ranged from 3.6 to 49.9 days^5,6,8,10,11,12,13^. Individual complication rates, like ventilator associated pneumonia or pressure sores were assessed in only a few studies, with largely varying results. We found only six studies^9,10,11,13,14,15^ that also discussed the prognostic factors associated with mechanical ventilation in ward. Individual studies used multivariate regression analysis, with the most consistent prognostic factors being higher APACHE-II or III score^9,11,13,14^, and recurrent or delayed intubation^10,15^.

Simchen et al^18^ had observed that admission into ICU has maximum benefit, compared to treatment in ward, in the initial three days after worsening of clinical status. In a study from Hong Kong by Joynt et al^19^, there was significantly higher standardized mortality ratios (SMRs) for those denied admission to an ICU (1.24; 1.05-1.46) as compared to those who were managed in ICU (0.93; 0.78-1.09), with Sinufu et al^20^ also demonstrating higher mortality among patients denied ICU admission (OR 3.04; 1.49-6.17), in a systematic review of ten observational studies. Sprung et al^21^, in a prospective observational study from Israel, showed a 14% mortality in patients admitted into the ICU, 36% mortality in those refused initially but admitted later and 46% among those patients who were never admitted. Therefore, it is a major responsibility to triage patients for admission into the ICU, for which, however, no universal admission criteria or validated scoring system is available.

Patients requiring prolonged mechanical ventilation are often shifted to wards to triage ICU beds, whose outcomes have rarely been studied. Few centres in Israel give preference to surgical patients in their post-operative period, where no separate surgical ICU exists, resulting in more medical mechanically ventilated cases being managed in wards^9^. Geriatric patients are denied ICU admission more frequently than young patients and have a higher ICU mortality when admitted, but the mortality benefit obtained by admitting in ICU vis-à-vis mechanical ventilation in ward is greater for the elderly, as per a multi-centre study by Sprung et al^22^. In the study by Hersch et al^9^ in Israel, young patients with lower day 1 APACHE score were preferentially admitted to ICU but multivariate analysis demonstrated that age was not an independent variable predicting death during hospitalization.

Triaging protocols and scores have not been properly validated for use in general medical wards, as was shown by our SR. Wongsurakiat^11^ showed that APACHE II score can be used to assess severity and predict outcome of critically ill patients admitted to general medical wards. Hersch et al^13^ showed that APS (Acute Physiology Score) part of APACHE III and Charlson index should be preferred over APACHE II for prognostication and management decisions. APS >90 was a predictor of increased mortality in non-ICU group of ventilated patients. Lieberman et al^14^ found significant differences in the functional status between intubated patients in ICU and non-ICU settings, with better pre-hospitalisation functional status among ICU patients. Non-ICU patients also had higher APACHE II scores. This is in accordance with the triaging practice in most hospitals across the globe whereby intubated patients with multiple comorbid illnesses and poor pre-hospitalisation functional status are preferably cared for in non-ICU settings.

A major limitation of the evidence is the paucity of literature on outcomes and prognostic factors of mechanical ventilation in the non-ICU (ward) setting, with existing studies been done only in a select few countries. As a result of this, our study demonstrated significant heterogeneity among individual studies both with respect to outcome variables like duration of hospital stay, duration of mechanical ventilation, mechanical ventilation related complications, and the prognostic factors associated with these outcomes. Also, the level and standard of care in non-ICU settings varies in each country/centre making generalisation of results difficult. The population studied was heterogeneous with better outcomes likely in patients who were being weaned after resolution of primary illness, or those who suffered traumatic injuries. The study by Iwashita et al^10^ had a much larger sample size compared to the other studies, significantly influencing the obtained results. Most other studies had small sample sizes and did not study all the outcome parameters and prognostic indicators. Nevertheless, this SR, which is the first such study on mechanical ventilation in non-ICU setting, highlights different notable aspects of the topic in question. It also reveals the dearth of evidence on several aspects, underlining the need for well-conceived, focused multi-centric studies over a substantial period of time.

## Supporting information

Study data for analysis

PRISMA 2020 checklist

PROSPERO registration protocol

ICMJE disclosure

## Data Availability

The data used in our systematic review and meta-analysis will be shared in the form of an excel (.xlsx) file.

## Acknowledgements/financial disclosures

None

## Conclusion

This systematic review and meta-analysis revealed that the cumulative incidence of mortality among patients receiving invasive mechanical ventilation in ward settings was high. Data regarding other outcomes and prognostic factors predicting mortality was very heterogeneous due to lack of suitable studies.

## Data availability

The data used in our systematic review and meta-analysis will be shared in a public repository as per the reviewers instructions.

## Bibliography

1. Hamid H, Abid Z, Amir A, Rehman TU, Akram W, Mehboob T. Current burden on healthcare systems in low-and middle-income countries: recommendations for emergency care of COVID-19. Drugs Ther Perspect. 2020 Aug 9:1–3.

2. Miller IF, Becker AD, Grenfell BT, Metcalf CJE. Disease and healthcare burden of COVID-19 in the United States. Nature Medicine. 2020 Aug;26(8):1212–7.

3. Kamio T, Masamune K. Mechanical Ventilation-Related Safety Incidents in General Care Wards and ICU Settings. Respir Care. 2018 Oct;63(10):1246–52.

4. Hui C, Lin M-C, Liu T-C, Wu R-G. Mortality and readmission among ventilator-dependent patients after successful weaned discharge from a respiratory care ward. J Formos Med Assoc. 2010 Jun;109(6):446–55.

5. Latriano B., McCauley P., Astiz M.E., Greenbaum D., Rackow E.C. Non-ICU care of hemodynamically stable mechanically ventilated patients. CHEST. 1996; 109(6): 1591–6.

6. Lin P.-H., Chen C.-F., Chiu H.-W., Lee D.L., Lai R.-S. Outcomes of unplanned extubation in ordinary ward are similar to those in intensive care unit: A retrospective case-control study. Respirology. 2018; 23:149.

7. DeBoisblanc M.W., Goldman R.K., Mayberry J.C., Brand D.M., Pangburn P.D., Soifer B.E., et al. Weaning injured patients with prolonged pulmonary failure from mechanical ventilation in a non-intensive care unit setting. J Trauma Inj Infect Crit Care. 2000;49(2):224–31.

8. Tang WM, Tong CK, Yu WC, Tong KL, Buckley TA. Outcome of adult critically ill patients mechanically ventilated on general medical wards. Hong Kong Med J. 2012 Aug;18(4):284–90.

9. Hersch M., Sonnenblick M., Karlic A., Einav S., Sprung C.L., Izbicki G. Mechanical ventilation of patients hospitalized in medical wards vs the intensive care unit-an observational, comparative study. J Crit Care. 2007;22(1):13–7.

10. Iwashita Y., Yamashita K., Ikai H., Sanui M., Imai H., Imanaka Y. Epidemiology of mechanically ventilated patients treated in ICU and non-ICU settings in Japan: a retrospective database study. Crit Care. 2018;22(1):329.

11. Wongsurakiat P., Sangsa N., Tangaroonsanti A. Mechanical ventilation of patients hospitalized on general medical ward: Outcomes and prognostic factors. J Med Assoc Thailand. 2016;99(7):764– 71.

12. Zisk-Rony R.Y., Weissman C., Weiss Y.G. Mechanical ventilation patterns and trends over 20 years in an Israeli hospital system: Policy ramifications. Isr J Health Policy Res. 2019;8(1).

13. Hersch M., Izbicki G., Dahan D., Breuer G.S., Nesher G., Einav S. Predictors of mortality of mechanically ventilated patients in internal medicine wards. J Crit Care. 2012;27(6):694–701.

14. Lieberman D, Nachshon L, Miloslavsky O, Dvorkin V, Shimoni A, Zelinger J, et al. Elderly patients undergoing mechanical ventilation in and out of intensive care units: a comparative, prospective study of 579 ventilations. Crit Care. 2010;14(2):R48.

15. Izhakian S, Buchs AE. Characterization of Patients who were Mechanically Ventilated in General Medicine Wards. Isr Med Assoc J. 2015 Aug;17(8):496–9.

16. Ambrosio IU, Woo MS, Jansen MT, Keens TG. Safety of hospitalized ventilator-dependent children outside of the intensive care unit. Pediatrics. 1998 Feb;101(2):257–9.

17. Wells G, Shea B, O’Connell D, Peterson je, Welch V, Losos M, et al. The Newcastle–Ottawa Scale (NOS) for Assessing the Quality of Non-Randomized Studies in Meta-Analysis. 2000 Jan 1;

18. Simchen E, Sprung CL, Galai N, Zitser-Gurevich Y, Bar-Lavi Y, Gurman G, et al. Survival of critically ill patients hospitalized in and out of intensive care units under paucity of intensive care unit beds. Crit Care Med 2004; 32: 1654–61.

19. Joynt GM, Gomersall CD, Tan P, Lee A, Cheng CA, Wong EL. Prospective evaluation of patients refused admission to an intensive care unit: triage, futility. Intensive Care Med 2001;27:1459–65.

20. Sinufuu T, Kahnamoui K, Cook DJ, et al. Rationing critical care beds: a systematic review. Crit Care Med. 2004;32:1588–97

21. Sprung CL, Geber D, Eidelman LA, Baras M, Pizov R, Nimrod A, et al. Evaluation of triage decisions for intensive care admission. Crit Care Med 1999; 27: 1073–9.

22. Sprung CL, Artigas A, Kesecioglu J, Pezzi A, Wiis J, Pirracchio R, et al. The Eldicus prospective, observational study of triage decision making in European intensive care units. Part II: intensive care benefit for the elderly. Crit Care Med 2012;40(1):132–8

