## Supplementary text and tables for "A study of clinical outcomes and prognostic factors associated with invasive mechanical ventilation of patients in non-ICU settings: A systematic review and meta-analysis"

### **Supplementary Appendix**

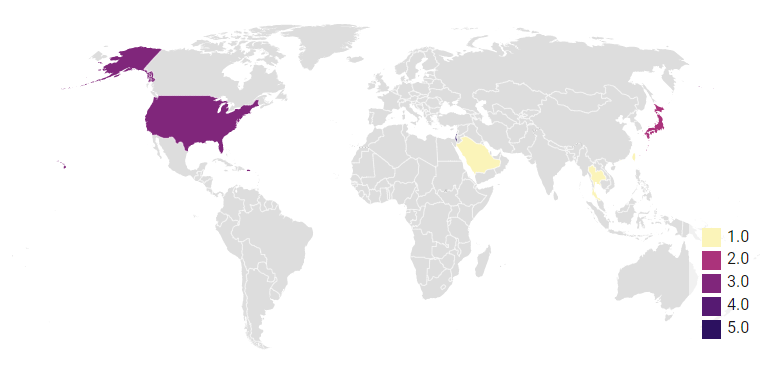

Figure 1: Global distribution of studies included in the Systematic Review

Table 1: Risk of bias in individual studies

| **Author, year** | **Selection** | | | | **Comparability^a^** | **Outcome** | | | **Total Score (out of 6)** | **Quality of study** |
| --- | --- | --- | --- | --- | --- | --- | --- | --- | --- | --- |
|  | 1 | 2^a^ | 3 | 4 | N/A | 1 | 2 | 3 |  |  |
| Hui et al, 2010^2^ | - | N/A | * | * | N/A | * | * | * | 5 | Good |
| Hersch M et al, 2007^7^ | * | N/A | * | * | N/A | * | * | * | 6 | Good |
| Wongsurakiat P et al, 2016^9^ | * | N/A | * | * | N/A | * | * | * | 6 | Good |
| Hersch M et al, 2012^11^ | * | N/A | * | * | N/A | * | * | * | 6 | Good |
| Lieberman et al, 2010^12^ | * | N/A | * | * | N/A | * | * | * | 6 | Good |
| Izhakian et al 2016^13^ | * | N/A | * | * | N/A | * | * | * | 6 | Good |

a- Not applicable (N/A) as all studies have only the exposed cohorts (no comparator)

**Newcastle Ottawa Scale subcomponents:**

**Selection:** 1: representativeness of exposed cohort; 2: selection of non-exposed cohort (N/A in our studies); 3: ascertainment of exposure; 4: demonstration that outcome of interest was not present at start of study

**Comparability:** N/A

**Outcome:** 1: assessment of outcome; 2: was follow up long enough for outcomes; 3: adequacy of follow-up

One star implies one point.

Original thresholds for converting the Newcastle-Ottawa scales to AHRQ standards (good, fair, and poor) with our modification:

**Good quality:** 3 or 4 stars in selection domain AND 1 or 2 stars in comparability domain AND 2 or 3 stars in outcome/exposure domain ()

**Fair quality:** 2 stars in selection domain AND 1 or 2 stars in comparability domain AND 2 or 3 stars in outcome/exposure domain

**Poor quality:** 0 or 1 star in selection domain OR 0 stars in comparability domain OR 0 or 1 stars in outcome/exposure domain

**Modifications:**

Comparability domain and selection of non-exposed cohort excluded.

**Good quality:** 2 or 3 stars in selection domain AND 2 or 3 stars in outcome/exposure domain

**Fair quality:** 1 star in selection domain AND 2 or 3 stars in outcome/exposure domain

**Poor quality:** 0 star in selection domain OR 0 or 1 stars in outcome/exposure domain

Table 2: Distribution of studies based on main parameter studied with respect to mechanical ventilation:

| **Parameter studied** | **Study** |
| --- | --- |
| Outcomes and prognostic factors | 7 studies^3,6,7,8,9,12,13^ |
| Ventilation related events | 2 studies^1,4^ |
| Weaning | 3 studies^2,5,14^ |
| Long term patterns and trends | 1 study^10^ |
| Predictors of mortality | 1 study^11^ |

Table 3: Baseline characteristics of patients

| Study population  Paediatric  Adult (≥ 18 y) | ICU  -  5 studies ^4,5,7,8,12^ |
| --- | --- |
| Males (%) | **ICU**  4 studies ^4,7,8,12^  12874/21053 (61.1%) |
| Diagnosis  Cardiovascular  Acute coronary syndrome  Pulmonary oedema/Heart failure  Arrythmia  Aortic dissection  Cardiogenic shock  Others/not specified  Neurological  Ischemic stroke  Intracranial bleed  Others/not specified  Respiratory cause  COPD/Asthma  ARDS  Respiratory failure  Pneumonia  -Interstitial  -Community acquired  -Hospital acquired  -Aspiration  -Others/not specified  Others/not specified  Sepsis (non-respiratory)  MODS  Post-operative  Post CPR  Trauma  Uraemia  Metabolic  Neoplastic  Drug overdose  Others/not specified | **ICU**  **5372 (25.8%)**  1340  2577  -  1424  15  16  **1640 (7.9%)**  4  1618  18  **2258 (10.8%)**  21  -  562  **1675**  462  479  11  691  -  32  1018 (4.9%)  -  10 (0.04%)  25 (0.1%)  7 (0.03%)  -  -  -  -  10513 (50.4%) |
|  | **20843** |
| Most common diagnosis overall  Neurological  Post-operative  Sepsis (non-pneumonic)  Respiratory  Heart failure | 1 study^7^  -  1 study^1^  -  1 study^8^ |

Table 4: Distribution of vital parameters (ward)

| **Variable** | **Study** | **Significant findings^*^** |
| --- | --- | --- |
| Blood pressure | Tang et al^6^ | Out of 426 patients, 755 (56.4%) were hypotensive pre- intubation. |
|  | Hersch et al^11^ | The mean of mean arterial pressure (MAP) of 86 patients in ward was 63.6±37.4 mmHg. MAP among survivors was 79.4±31.6 mmHg as compared to 58.2±37.9 mmHg among non-survivors. |
|  | Izhakian et al^13^ | Out of 437 patients, mean systolic blood pressure (SBP) among survivors was 134.8±26.2 mmHg, as compared to 121.5±30.4 mmHg among non-survivors. Mean diastolic blood pressure (DBP) among survivors was 73.6±16.3 mmHg, as compared to 67.2±17.9 mmHg among non-survivors. |

^*^Findings are statistically significant in individual studies (p values<0.05)

Table 5: Investigations (ward)

| **Variable** | **Study** | **Significant findings^*^** |
| --- | --- | --- |
| Serum albumin | Latriano et al^3^ | Among 224 patients in the respiratory care ward, mean serum albumin was 2.78±0.49 g/dL. Mean albumin was 2.9±0.5 g/dL among survivors as compared to 2.6±0.4 g/dL among non-survivors. |
|  | Hersch et al^7^ | Mean albumin among patients in ward (n=65) was 3.5±0.3 g/dL, as compared to 3.3±0.8 in ICU (n=34). |
|  | Hersch et al^11^ | Among 86 patients ventilated in ward, mean serum albumin among survivors was 2.98±0.66 g/dL as compared to 2.67±0.73 g/dL among non-survivors. |
| Renal dysfunction | Hersch et al^11^ | Mean serum creatinine was 1.95±1.5 mg/dL, with a value of 1.2±0.9 mg/dL among survivors as compared to 2.3±1.6 mg/dL among non-survivors.  Mean urine output was 1389±1155 mL/day, with 2035±1112 mL/day among survivors and 1066±1052 mL/day among non-survivors. |
|  | Izhakian et al^13^ | Among 437 patients, mean serum creatinine among survivors was 1.3±0.8 mg/dL, as compared to 1.8±1.5 mg/dL among non-survivors. |

^*^Findings are statistically significant in individual studies (p values<0.05)

Table 6: Monitoring and treatment (ward)

| **Variable** | **Study** | **Significant findings (ward)^*^** |
| --- | --- | --- |
| Ventilatory changes | Hersch et al^7^ | An average of 1.3±1.0 ventilatory changes were made in a day. |
| Arterial blood gas (ABG) analysis and invasive monitoring | Hersch et al^7^ | On an average, 2.3±1.3 ABGs were done in a day. |
|  | Iwashita et al^8^ | Out of 17775 patients in ward, arterial line was present in 3057 (17.2%) patients and central line was inserted for 7412 (41.7%) patients. |
| Use of vasopressors, renal replacement therapy, stress ulcer prophylaxis, sedation and neuromuscular blockade | Iwashita et al^8^ | In ward, vasopressor use was seen in 7945 patients (44.7%). Renal replacement therapy was used in 1404 patients (7.9%). Sedation was used in 7234 patients (40.7%) and opioids were used in 2453 patients (13.8%). Neuromuscular blockade for > 48 hours was done in 462 patients (2.6%). Stress ulcer prophylaxis for >72 hours, was given to 6470 patients (36.4%). |

^*^Findings are statistically significant in individual studies (p values<0.05)

Table 7: Mechanical ventilation related events:

| **Variable** | **Study** | **Significant findings^*^** |
| --- | --- | --- |
| Ventilator related adverse events | Kamio et al^1^ | There were 182 events in ward; 6 (3%) ventilator malfunctions, 7 (4%) equipment damages, 72 (40%) airway or breathing circuit issues, 1 power source issue and 96 (53%) human factor issues. There were 79 events in the ICU, with no significant difference in the rate of different events. |
|  | Hersch et al^7^ | Endotracheal tube related events were seen in 40 out of 65 patients (62%) in the ward. |
| Unplanned extubations | Lin et al^4^ | In ward, there were 102 unplanned extubations (88 self- and 14 accidental extubations). |
|  | Tang et al^6^ | Out of 755 mechanically ventilated patients, 19 (2.5%) had unplanned extubations in general ward. |

^*^Findings are statistically significant in individual studies (p values<0.05)

Table 8: Comparison of significant clinical, monitoring and treatment parameters between ward and ICU

| **Variable** | **Study** | **Significant findings^*^** |
| --- | --- | --- |
| Prognostic scores | Hersch et al^7^  Lieberman et al^12^ | Mean APACHE II score was significantly higher in ward (27±7) compared to ICU (24±7).  Mean acute physiology score (APS) in ward (15±5) was significantly higher than ICU (13.8±6.4). |
| Functional status (FIM score) | Lieberman et al^12^ | Decision to ventilate in ICUs was independently influenced by pre-hospitalization functional status, assessed by functional independence measure (FIM) scale for motor function and cognition. The average total FIM score, motor and cognitive FIM scores in the ICU cohort were 104.9 ± 19.5, 73.4±16.7 and 31.5±4.4 respectively, and were significantly higher than the non-ICU cohort, which were 75.8 ± 31.2, 51.3±24.1 and 24.3±8.8, respectively. |
| Unplanned extubations | Lin et al^4^ | Out of 9245 total intubated patients, there were 102 (44.4%) unplanned extubations in non-ICU settings (88 self- and 14 accidental extubations), while in ICU there were 220 (31.6%) unplanned extubations (211 self and 9 accidental extubations). Admission in non-ICU setting was significantly associated with unplanned extubations. |
| Endotracheal tube related events | Hersch et al^7^ | Endotracheal tube related events were significantly more common in ward, seen in 40 out of 65 patients (62%), as compared to ICU, with events in 7 out of 34 (20%) patients. |
| Ventilatory changes | Hersch et al^7^ | In ward, an average of 1.3±1.0 ventilatory changes were made in a day, compared to significantly more (7.5±1.4) changes in the ICU. |
| Arterial blood gas (ABG) analysis and invasive monitoring | Hersch et al^7^ | In ward, an average of 2.3±1.3 ABGs were done in a day, compared to 7.7±1.2 in the ICU, which was significantly higher. This was probably because 97% patients in ICU had an arterial line, but none of the ward patients had one. |
|  | Iwashita et al^8^ | Arterial line was present in 3057 out of 17775 patients (17.2%) in ward and 13294 out of 20516 patients (64.8%) in ICU. A central line was inserted for 7412 patients (41.7%) as compared to 13335 (65%) in the ICU. |
| Use of vasopressors, renal replacement therapy, stress ulcer prophylaxis sedation and neuromuscular blockade | Iwashita et al^8^ | Vasopressor use was seen in 7945 patients (44.7%) in ward as compared to 13992 patients (68.2%) in ICU. Use of renal replacement therapy was seen in 1404 patients (7.9%) in ward vs 3488 patients (17%) in the ICU. Use of sedation was seen in 7234 patients (40.7%) in ward, as compared to 14443 patients (70.4%) in ICU. Opioid use was seen in 2453 patients (13.8%) in ward vs 8227 (40.1%) in the ICU. Neuromuscular blockade for > 48 hours was seen in 462 patients (2.6%) in ward, compared to 1436 (7%) in the ICU. Stress ulcer prophylaxis for > 72 hours was given to 6470 patients (36.4%) in ward vs 12309 patients (60%) in ICU. All parameters were significantly higher in the ICU cohort. |
| Duration of hospital stay/mechanical ventilation | Iwashita et al^8^ | Total duration of mechanical ventilation was significantly higher in non-ICU settings, while duration of hospital stay was significantly higher in the ICU cohort. Duration of ventilation was 11.7 days in ward compared to 9.5 days in ICU, while duration of hospital stay was 24.7 days in ward, compared to 26.1 days in ICU group. |

^*^Findings are statistically significant in individual studies (p values<0.05)

Table 9: Reasons for heterogeneity in studies excluded from meta-analysis

| **Study** | **Mortality rate** | **Reason for heterogeneity** |
| --- | --- | --- |
| Latriano et al^3^ | 49.6% | For mechanical ventilation in ward, a dedicated respiratory care floor (RCF) was delineated, where intensivists, respiratory therapists, and nurses provided care, apart from the doctors in ward.  Only hemodynamically stable patients, not requiring vasoactive drug infusions for previous 24 h, with an FiO2 requirement of ≤50% and PEEP requirement ≤ 10 cm of H_2_O, were transferred to the RCF.  Superior standard of care on the RCF as compared to ward, and more stable patient profile, were responsible for the lower mortality in this study. |
| Lin et al^4^ | 28 deaths | Lin reported total number of deaths in ward and ICU, and the number of events in ward and ICU. Although the total number of patients studied has been mentioned, the number of patients separately in ward and ICU have not been mentioned in the study, and hence estimation of mortality rates was not possible. |
| deBoisblanc et al^5^ | 3.9% | Post-trauma hemodynamically stable patients requiring weaning, had a tracheostomy and required mechanical ventilation > 7 days were included. Patients had a spontaneous respiratory drive and required low levels of ventilatory support through pressure support ventilation (PSV) with pressure support of 15 cm H2O and PEEP of 5 cm H2O and FiO2 requirement of ≤ 0.5. After adaptation to PSV for 24 h in ICU, they were then shifted to ward.  This cohort had significantly lower mortality as only trauma patients whose primary condition had resolved, and required weaning were included. |
| Tang et al^6^ | 89.1% | Patients on mechanical ventilation who were too sick and unlikely to benefit from ICU care were managed in wards. Patients in respiratory care wards were excluded.  307/755 (40.7%) of the patients were on invasive mechanical ventilation after cardiopulmonary resuscitation for cardiac arrest. This large sub-cohort had a mortality rate of 99%, which was significantly higher than overall mortality (P<0.0005)  Thus, mortality in this study was significantly higher. |
| Iwashita et al^8^ | 41.4% | ‘Quasi-ICU units’ treating severely ill patients and functioning like ICUs were counted among non-ICUs in this study, and thus had a much better level of care.  Patients with poor prognosis, like cancer patients, and those with hospital stay > 60 days, were excluded. |
