## Supplementary material for "A study of clinical outcomes and prognostic factors associated with invasive mechanical ventilation of patients in non-ICU settings: A systematic review and meta-analysis": PROSPERO registration protocol

### Systematic review

#### 1. * Review title.

Give the working title of the review, for example the one used for obtaining funding. Ideally the title should state succinctly the interventions or exposures being reviewed and the associated health or social problems. Where appropriate, the title should use the PI(E)COS structure to contain information on the Participants, Intervention (or Exposure) and Comparison groups, the Outcomes to be measured and Study designs to be included.

A study of clinical outcomes and prognostic factors associated with invasive mechanical ventilation of adult patients in non ICU settings: A systematic review

#### 2. Original language title.

For reviews in languages other than English, this field should be used to enter the title in the language of the review. This will be displayed together with the English language title.

#### 3. * Anticipated or actual start date.

Give the date when the systematic review commenced, or is expected to commence.

17/01/2020

#### 4. * Anticipated completion date.

Give the date by which the review is expected to be completed.

17/04/2020

#### 5. * Stage of review at time of this submission.

Indicate the stage of progress of the review by ticking the relevant Started and Completed boxes. Additional information may be added in the free text box provided.
Please note: Reviews that have progressed beyond the point of completing data extraction at the time of initial registration are not eligible for inclusion in PROSPERO. Should evidence of incorrect status and/or completion date being supplied at the time of submission come to light, the content of the PROSPERO record will be removed leaving only the title and named contact details and a statement that inaccuracies in the stage of the review date had been identified.
This field should be updated when any amendments are made to a published record and on completion and publication of the review. If this field was pre-populated from the initial screening questions then you are not able to edit it until the record is published.

The review has not yet started: No

| Review stage | Started | Completed |
|---|---|---|
| Preliminary searches | Yes | Yes |
| Piloting of the study selection process | Yes | No |
| Formal screening of search results against eligibility criteria | Yes | No |
| Data extraction | No | No |
| Risk of bias (quality) assessment | No | No |
| Data analysis | No | No |

Provide any other relevant information about the stage of the review here (e.g. Funded proposal, protocol not yet finalised).

#### 6. * Named contact.

The named contact acts as the guarantor for the accuracy of the information presented in the register record.

Animesh Ray

#### Email salutation (e.g. "Dr Smith" or "Joanne") for correspondence:

Dr Ray

#### 7. * Named contact email.

Give the electronic mail address of the named contact.

#### 8. Named contact address

Give the full postal address for the named contact.

#### 9. Named contact phone number.

Give the telephone number for the named contact, including international dialling code.

+91-9560093190

#### 10. * Organisational affiliation of the review.

Full title of the organisational affiliations for this review and website address if available. This field may be completed as 'None' if the review is not affiliated to any organisation.

All India Institute of Medical Sciences

#### Organisation web address:

#### 11. * Review team members and their organisational affiliations.

Give the personal details and the organisational affiliations of each member of the review team. Affiliation refers to groups or organisations to which review team members belong. **NOTE: email and country are now mandatory fields for each person.**

Dr Shubham Agarwal. All India Institute of Medical Sciences
Dr Animesh Ray. All India Institute of Medical Sciences
Dr Abhishek Anand. All India Institute of Medical Sciences
Dr Ranveer Jadon. All India Institute of Medical Sciences
Dr Naval Vikram. All India Institute of Medical Sciences
Dr Ananthu Narayan. All India Institute of Medical Sciences
Dr Neha Chopra. All India Institute of Medical Sciences
Dr Vishakh Kheri. All India Institute of Medical Sciences
Dr Neha Rastogi. All India Institute of Medical Sciences
Dr Debarchan Roy. All India Institute of Medical Sciences
Dr Aishwarya Ramprasad. All India Institute of Medical Sciences

#### 12. * Funding sources/sponsors.

Give details of the individuals, organizations, groups or other legal entities who take responsibility for initiating, managing, sponsoring and/or financing the review. Include any unique identification numbers assigned to the review by the individuals or bodies listed.

None

#### Grant number(s)

#### 13. * Conflicts of interest.

List any conditions that could lead to actual or perceived undue influence on judgements concerning the main topic investigated in the review.

None

#### 14. Collaborators.

Give the name and affiliation of any individuals or organisations who are working on the review but who are not listed as review team members. **NOTE: email and country are now mandatory fields for each person.**

#### 15. * Review question.

State the question(s) to be addressed by the review, clearly and precisely. Review questions may be specific or broad. It may be appropriate to break very broad questions down into a series of related more specific questions. Questions may be framed or refined using PI(E)COS where relevant.

To review various clinical outcomes, mortality rates and prognostic factors associated with invasive

mechanical ventilation of adult patients for 48 hours, in non ICU (ward) settings

#### 16. * Searches.

State the sources that will be searched. Give the search dates, and any restrictions (e.g. language or publication period). Do NOT enter the full search strategy (it may be provided as a link or attachment.)

PubMed, Embase, Indmed (current to 1970)

#### 17. URL to search strategy.

Give a link to a published pdf/word document detailing either the search strategy or an example of a search strategy for a specific database if available (including the keywords that will be used in the search strategies), or upload your search strategy.Do NOT provide links to your search results.

Alternatively, upload your search strategy to CRD in pdf format. Please note that by doing so you are consenting to the file being made publicly accessible.

Do not make this file publicly available until the review is complete

#### 18. * Condition or domain being studied.

Give a short description of the disease, condition or healthcare domain being studied. This could include health and wellbeing outcomes.

Outcomes of mechanical ventilation in ICUs are well known but less is known about clinical and mortality outcomes at centres where invasive mechanical ventilation is done in a non ICU (ward) setting. At resource limited centres, managing such sick patients in wards is a necessity but we do not know how effective it is, without having robust literature on outcomes and prognostic factors in this setting.

#### 19. * Participants/population.

Give summary criteria for the participants or populations being studied by the review. The preferred format includes details of both inclusion and exclusion criteria.

All articles (barring editorial reviews and narrrative reviews) reporting mechanical ventilation done in a non ICU/ward setting.

#### 20. * Intervention(s), exposure(s).

Give full and clear descriptions or definitions of the nature of the interventions or the exposures to be reviewed.

Invasive mechanical ventilation

#### 21. * Comparator(s)/control.

Where relevant, give details of the alternatives against which the main subject/topic of the review will be compared (e.g. another intervention or a non-exposed control group). The preferred format includes details of both inclusion and exclusion criteria.

none

#### 22. * Types of study to be included.

Give details of the types of study (study designs) eligible for inclusion in the review. If there are no restrictions on the types of study design eligible for inclusion, or certain study types are excluded, this should be stated. The preferred format includes details of both inclusion and exclusion criteria.

prospective/retrospective/ambispective observational studies

#### 23. Context.

Give summary details of the setting and other relevant characteristics which help define the inclusion or exclusion criteria.

Invasive mechanical ventilation in a non ICU/ward setting exclusively, among adults, for 48 hours.

#### 24. * Main outcome(s).

Give the pre-specified main (most important) outcomes of the review, including details of how the outcome is defined and measured and when these measurement are made, if these are part of the review inclusion criteria.

mortality rates

##### * Measures of effect

Please specify the effect measure(s) for you main outcome(s) e.g. relative risks, odds ratios, risk difference, and/or 'number needed to treat.

none

#### 25. * Additional outcome(s).

List the pre-specified additional outcomes of the review, with a similar level of detail to that required for main outcomes. Where there are no additional outcomes please state ‘None’ or ‘Not applicable’ as appropriate to the review

Clinical outcomes like physiological scores (APACHE, SAPS), prognostic factors

##### * Measures of effect

Please specify the effect measure(s) for you additional outcome(s) e.g. relative risks, odds ratios, risk difference, and/or 'number needed to treat.

none

#### 26. * Data extraction (selection and coding).

Describe how studies will be selected for inclusion. State what data will be extracted or obtained. State how this will be done and recorded.

The total number of researchers are going to be 11. Five of them are going to individually look for
Studies having details of invasive mechanical ventilation in the ICU/ward setting and five researchers will be double-checking and proof-reading their work. All disagreements will be sorted out by Professor Vikram (Head). All the studies meeting the eligibility criteria and indexed in PubMed, Embase or Indmed and whose abstract/full text contains the above details will be included in the systematic review. Studies not meeting the above criteria would not be included in the review.

Information about study design, participant demographics and baseline characteristics, numbers of events (mortalities) and clinical physiological scores (APACHE, SAPS), prognostic factors will be extracted. The data will be extracted by all 11 researchers. Study investigators will not be contacted for providing missing data. All data will be recorded in an Excel sheet.

#### 27. * Risk of bias (quality) assessment.

Describe the method of assessing risk of bias or quality assessment. State which characteristics of the studies will be assessed and any formal risk of bias tools that will be used.

Assessment will be done at study and outcome level

Cochrane risk of bias tool will be used for assessment

5 reviewers will be involved in the quality assessment

#### 28. * Strategy for data synthesis.

Provide details of the planned synthesis including a rationale for the methods selected. This **must not be**

**generic text** but should be **specific to your review** and describe how the proposed analysis will be applied to your data.

Minimum 3 studies would be required. Outcomes like event rates will be noted. Aggregate participant data will be used.

#### 29. * Analysis of subgroups or subsets.

State any planned investigation of 'subgroups'. Be clear and specific about which type of study or participant will be included in each group or covariate investigated. State the planned analytic approach.

No subgroup analysis

#### 30. * Type and method of review.

Select the type of review and the review method from the lists below. Select the health area(s) of interest for your review.

##### Type of review

Cost effectiveness
No

Diagnostic
No

Epidemiologic
No

Individual patient data (IPD) meta-analysis
No

Intervention
Yes

Meta-analysis
No

Methodology
No

Narrative synthesis
No

Network meta-analysis
No

Pre-clinical
No

Prevention
No

Prognostic
No

Prospective meta-analysis (PMA)
No

Review of reviews
No

Service delivery
No

Synthesis of qualitative studies
No

Systematic review
Yes

Other

No

#### Health area of the review

Alcohol/substance misuse/abuse
No

Blood and immune system
No

Cancer
No

Cardiovascular
No

Care of the elderly
No

Child health
No

Complementary therapies
No

Crime and justice
No

Dental
No

Digestive system
No

Ear, nose and throat
No

Education
No

Endocrine and metabolic disorders
No

Eye disorders
No

General interest
No

Genetics
No

Health inequalities/health equity
No

Infections and infestations
No

International development
No

Mental health and behavioural conditions
No

Musculoskeletal
No

Neurological
No

Nursing
No

Obstetrics and gynaecology

No

Oral health
No

Palliative care
No

Perioperative care
No

Physiotherapy
No

Pregnancy and childbirth
No

Public health (including social determinants of health)
No

Rehabilitation
No

Respiratory disorders
No

Service delivery
No

Skin disorders
No

Social care
No

Surgery
No

Tropical Medicine
No

Urological
No

Wounds, injuries and accidents
No

Violence and abuse
No

#### 31. Language.

Select each language individually to add it to the list below, use the bin icon  to remove any added in error.
English

There is not an English language summary

#### 32. * Country.

Select the country in which the review is being carried out from the drop down list. For multi-national collaborations select all the countries involved.

India

#### 33. Other registration details.

Give the name of any organisation where the systematic review title or protocol is registered (such as with The Campbell Collaboration, or The Joanna Briggs Institute) together with any unique identification number assigned. (N.B. Registration details for Cochrane protocols will be automatically entered). If extracted data will be stored and made available through a repository such as the Systematic Review Data Repository (SRDR), details and a link should be included here. If none, leave blank.

none

#### 34. Reference and/or URL for published protocol.

Give the citation and link for the published protocol, if there is one

Give the link to the published protocol.

Alternatively, upload your published protocol to CRD in pdf format. Please note that by doing so you are consenting to the file being made publicly accessible.

No I do not make this file publicly available until the review is complete

Please note that the information required in the PROSPERO registration form must be completed in full even if access to a protocol is given.

#### 35. Dissemination plans.

Give brief details of plans for communicating essential messages from the review to the appropriate audiences.

#### Do you intend to publish the review on completion?

Yes

#### 36. Keywords.

Give words or phrases that best describe the review. Separate keywords with a semicolon or new line. Keywords will help users find the review in the Register (the words do not appear in the public record but are included in searches). Be as specific and precise as possible. Avoid acronyms and abbreviations unless these are in wide use.

Mechanical ventilation, ward, APACHE score

#### 37. Details of any existing review of the same topic by the same authors.

Give details of earlier versions of the systematic review if an update of an existing review is being registered, including full bibliographic reference if possible.

#### 38. * Current review status.

Review status should be updated when the review is completed and when it is published. For newregistrations the review must be Ongoing.
Please provide anticipated publication date

Review_Ongoing

#### 39. Any additional information.

Provide any other information the review team feel is relevant to the registration of the review.

#### 40. Details of final report/publication(s).

This field should be left empty until details of the completed review are available.

Give the link to the published review.
